# Comprehensive analysis of the key epidemiological parameters to evaluate the impact of BCG vaccination on COVID-19 pandemic

**DOI:** 10.1101/2020.08.12.20173617

**Authors:** Niloy R. Datta, Sneha Datta

## Abstract

Globally, the heterogenous coronavirus disease 2019 (COVID-19) case fatality rate (CFR) could be influenced by various epidemiological parameters. Identifying these could help formulate effective public health strategies. Incidence and mortality of COVID-19 for each of the 220 countries as on July 30, 2020 were evaluated against key epidemiological variables, namely - BCG vaccination (ongoing vs. discontinued/never undertaken), %population aged ≥65 years, incidences of ischemic heart disease (IHD), hypertensive heart disease (HHD), cancer, malaria, and diabetes; human development index (HDI) and population density. These were retrieved from the public domains of WHO, UN, World Bank and published reports. The COVID-19 CFRs ranged between 0.0% and 28.3% (mean ± SD: 3.05% ± 3.48). The influence of the individual epidemiological parameters on CFR were evaluated through the event rate estimations. A significantly lower event rate was observed in countries with ongoing BCG vaccination program (ER: with vs without ongoing BCG vaccination: 0.020 vs 0.034, p<0.001). The type of BCG strains used also influenced the ER; this being 0.018, 0.031 and 0.019 for early, late and mixed strains respectively (p=0.008). The epidemiological variables significantly associated with higher COVID-19 event rate were countries with higher %population aged ≥65 years (p<0.001), greater incidence of IHD (p<0.001) and cancer (p=0.003) and better HDI (p=0.003). Incidences of malaria, HHD and diabetes along with population density had no significant impact on COVID-19 CFR. Further, BCG vaccination significantly lowered the COVID-19 ER in each of the high-risk population subgroups - countries with >7.1% population aged ≥65 years (p=0.008), >0.737 HDI (p=0.001), IHD >1171/10^5^ population (p=0.004) and cancer incidence >15726 (p<0.001). The results supports BCG induced “trained immunity” leading to heterologous immunoprotection against COVID-19. Thus BCG vaccination with early strains could provide a cost-effective prophylaxis, especially in high-risk individuals and bridge the gap till an effective vaccine against SARS-CoV-2 is freely available globally.

## Introduction

As the global community eagerly waits for a successful vaccine against severe acute respiratory sickness coronavirus 2 (SARS-CoV-2) responsible for the ongoing pandemic of coronavirus disease 2019 (COVID-19), the morbidity and mortality continues to rise unabated. While some countries have seen off the passing of the first wave of COVID-19, others continue to face the onslaught with a rising trend. Further, the danger of a second wave still looms large over the regions that have contained the first wave of COVID-19 with varying degrees of morbidity and mortality.

A glance at the world-wide pattern of case fatality of COVID-19 reveals a gross heterogeneity in its distribution. This could be attributed to a complex interaction of host factors that could include the virulence of the strains of SARS-CoV-2, appropriate enforcement of containment measures, screening and contact tracing, population density, availability of adequate medical care, inherent susceptibility of the population, associated comorbid conditions, socioeconomic status and others [1-9]. Presently, although there are some indications of the individual factors to impact the COVID-19 mortality, a comprehensive evaluation of the relevant epidemiological variables towards the heterogenous case fatality is needed to identify parameter/s that might help to alleviate the mortality and consider active interventions in high-risk population.

Thus, a comprehensive analysis has been carried out by estimating the event rates to evaluate the influence of key epidemiological factors on the case fatality rate (CFR) of COVID-19. As the policy of subjecting the inhabitants of a country to routine BCG vaccination is the variable in the 220 countries evaluated, the purpose of this study is to evaluate the likely impact of BCG vaccination on the heterogenous COVID-19 CFR. Thus the effect of BCG vaccination and the strains used have been studied to specifically estimate its impact on each of the high-risk populations as evident from this study. This might assist the national health agencies to formulate country specific public health guidelines to mitigate the COVID-19 case fatality, especially in high-risk inhabitants.

## Methods

### Databases used

The estimates of total cases, total deaths, cases/million and deaths/million due to COVID-19 were extracted for each country and their dependent territories as listed in the WHO [10] and Worldometer [11] websites as on July 7, 2020 (Fig. 1). COVID-19 CFR for each country was computed using the total cases and deaths listed as (total deaths/total cases) x 100.

**Fig. 1.**
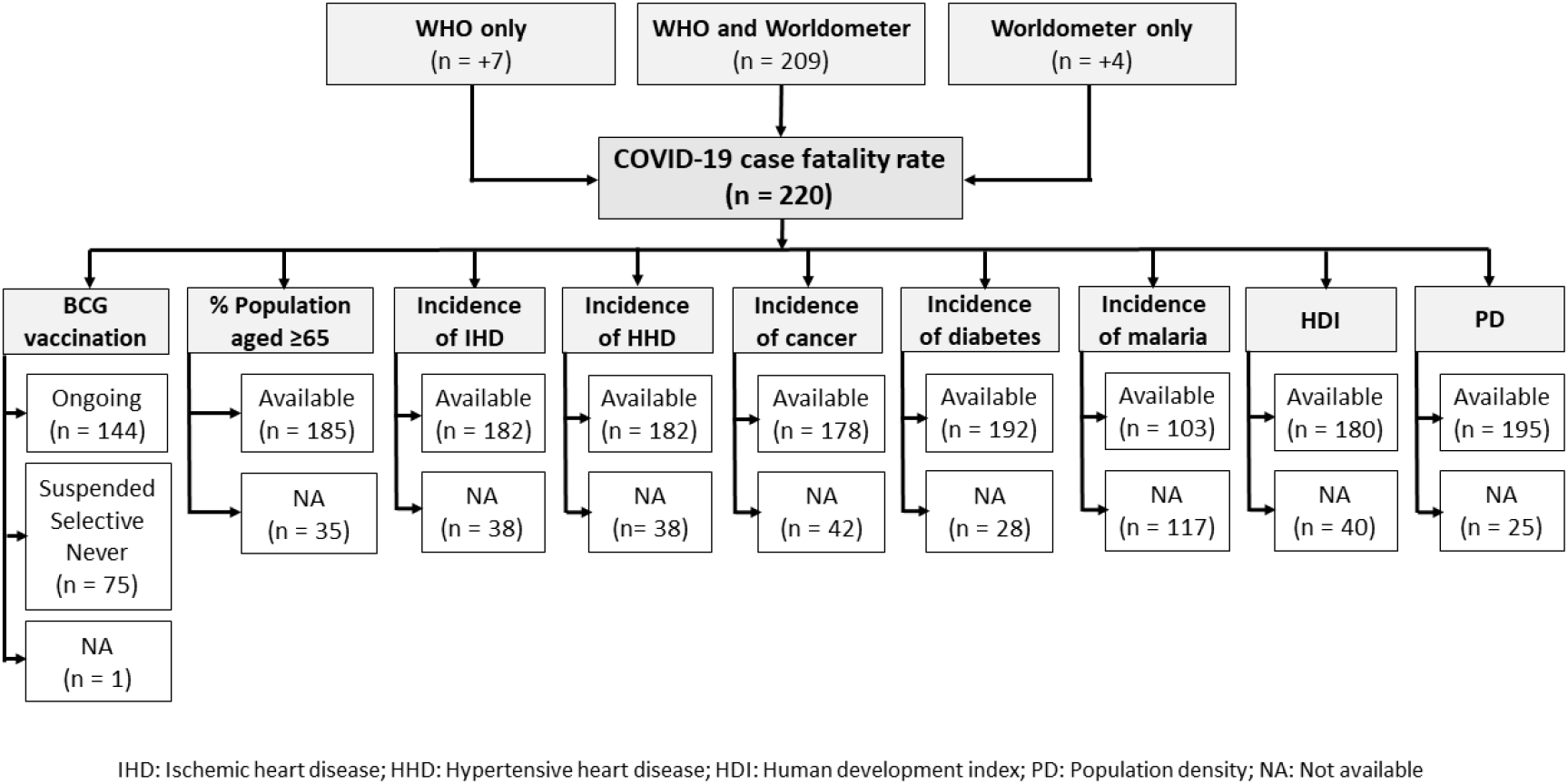
Flow chart indicating the sources of COVID-19 case fatality and the availability of the data for the relevant epidemiological parameters retrieved from the public domains of various websites. The details of the websites from where the data have been extracted are mentioned in the text.

The epidemiological parameters were retrieved from the public domains of WHO, UN, World Bank, BCG World Atlas and published reports for each listed countries (Fig. 1). These included - %BCG vaccination (2018) [12, 13], %population aged ≥65 years (2018) [14], incidences of ischemic heart disease (IHD) (2015) [15], hypertensive heart disease (HHD) (2015) [15], cancer (2020) [16], malaria (2018) [17] and diabetes [18]; human development index (HDI) (2019) [19] and population density (2018) [20]. In all cases, the latest values listed in the above public domains were considered.

Based on the national BCG immunization program, the countries were grouped as, either (a) countries where the BCG vaccination is currently not ongoing or those where is had been discontinued or in those where its recommended selectively for the high-risk groups (e.g. health workers) and (b) countries with ongoing BCG program for all its inhabitants.

It was also evident that BCG strains used for their immunization program varied in these countries. Broadly these countries are classified as using early (BCG Russia, Brazil, Japan, Sweden, Birkhaug) or late BCG strains (BCG Prague, China, Glaxo, Denmark, Tice, Frappier, Connaught, Phipps, Pasteur) [21, 22], Some countries had also used mixed BCG strains. The details of the type of BCG strain used for each of the countries with ongoing BCG vaccination was retrieved and collated from BCG World Atlas [13] and also the published reports [21, 22] and classified as early, late and mixed BCG strains.

### Grouping based on case fatality rates

Based on their COVID-19 CFR, the 220 countries were divided into low, moderate and high risk groups. The cutoff values were determined as (a) low risk group with CFR ≤33^rd^ percentile (b) moderate risk group with CFR values between 34^th^ and 66^th^ percentile and (c) high risk group with CFR >66^th^ percentile. The key epidemiological variables were estimated and the prognostic impact of these on the CFR for each of the 3 groups were evaluated.

### Statistical considerations: COVID-19 event rate and case fatality rate analysis

COVID-19 events rates for each of the 220 countries were computed as events/nonevents. For each country, the events indicate the number of deaths, while nonevents were the difference between the number of confirmed cases and resulting deaths. Comprehensive Meta-analysis Software (version 3.0) was used to perform the event rate analysis [23].

The event rates for the three subgroups of BCG strains were compared using mixed effects model. The median value for each of the parameters namely - %population aged ≥65 years, incidences of IHD, HHD, cancer, malaria and diabetes; HDI and population density were used to create two distinct groups for each of these epidemiological covariate.

BCG vaccination was considered as a categorical variable based on the presence or absence of ongoing BCG vaccination program in each country. The effect of BCG vaccination have been studied to specifically estimate its impact on the high-risk groups of the other epidemiological variables. Comparative subgroup analysis for each of the above variables were carried out using mixed effects model and forest plots were generated along with point estimates, 95% confidence intervals, Q value, I^2^ and p value.

For CFR, significance of comparison between two groups were reported using Mann-Whitney U test while for 3 groups, Kruskal-Wallis test [24] was used.

## Results

### COVID-19: Case fatality rate

Of the 220 countries and their dependent territories, 209 were represented in both the WHO and Worldometer websites while seven additional countries were extracted from the WHO [10] and four from Worldometer [11] public domains (Fig.1). As of July 30, 2020, a total of 17.3 million cases of COVID-19 have been reported from 220 countries worldwide (S1 Table). The total cases reported from these countries have ranged from 3 to 4.58 million (mean ± SD: 78716.5 ± 377563). A total of 0.67 million deaths have been ascribed to COVID-19 and this varied between 0 to 0.15 million in these 220 countries. Consequently the COVID-19 CFR ranged from 0% to 28.3% (median: 2.1%) (Fig.2a). The gross heterogeneity in the cases/million, deaths/million and COVID-19 CFR are evident from Table 1.

**Fig. 2.**
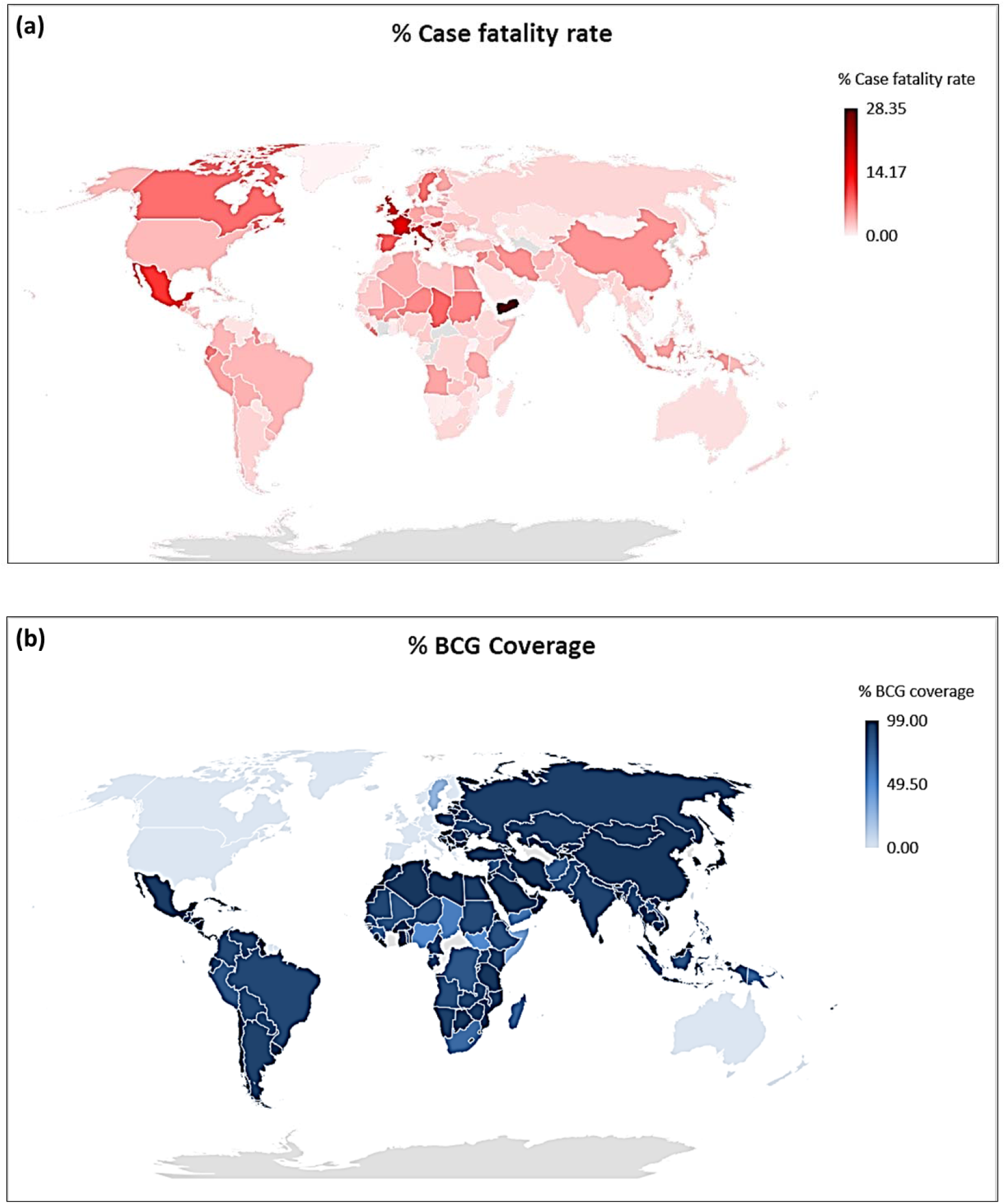
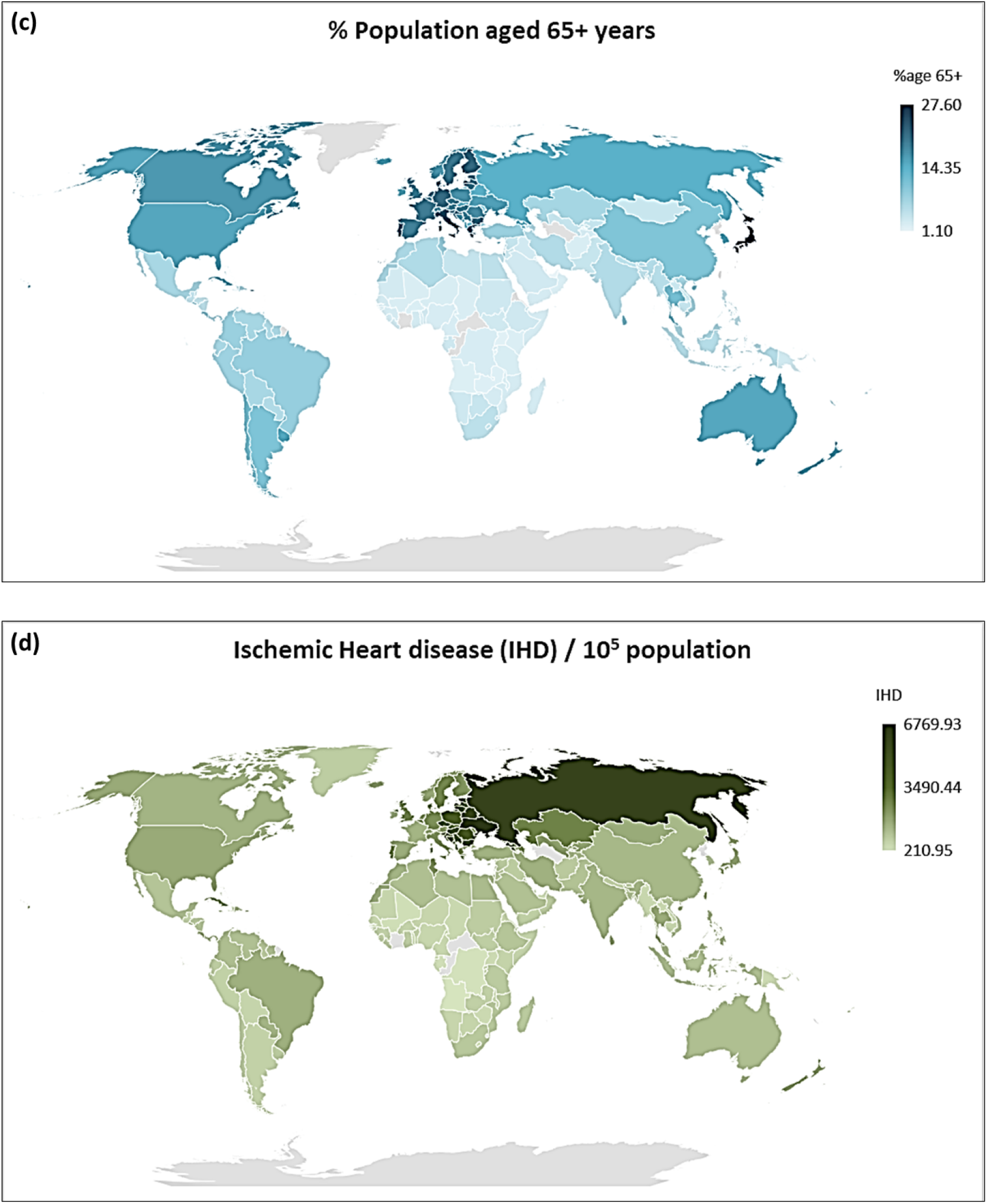
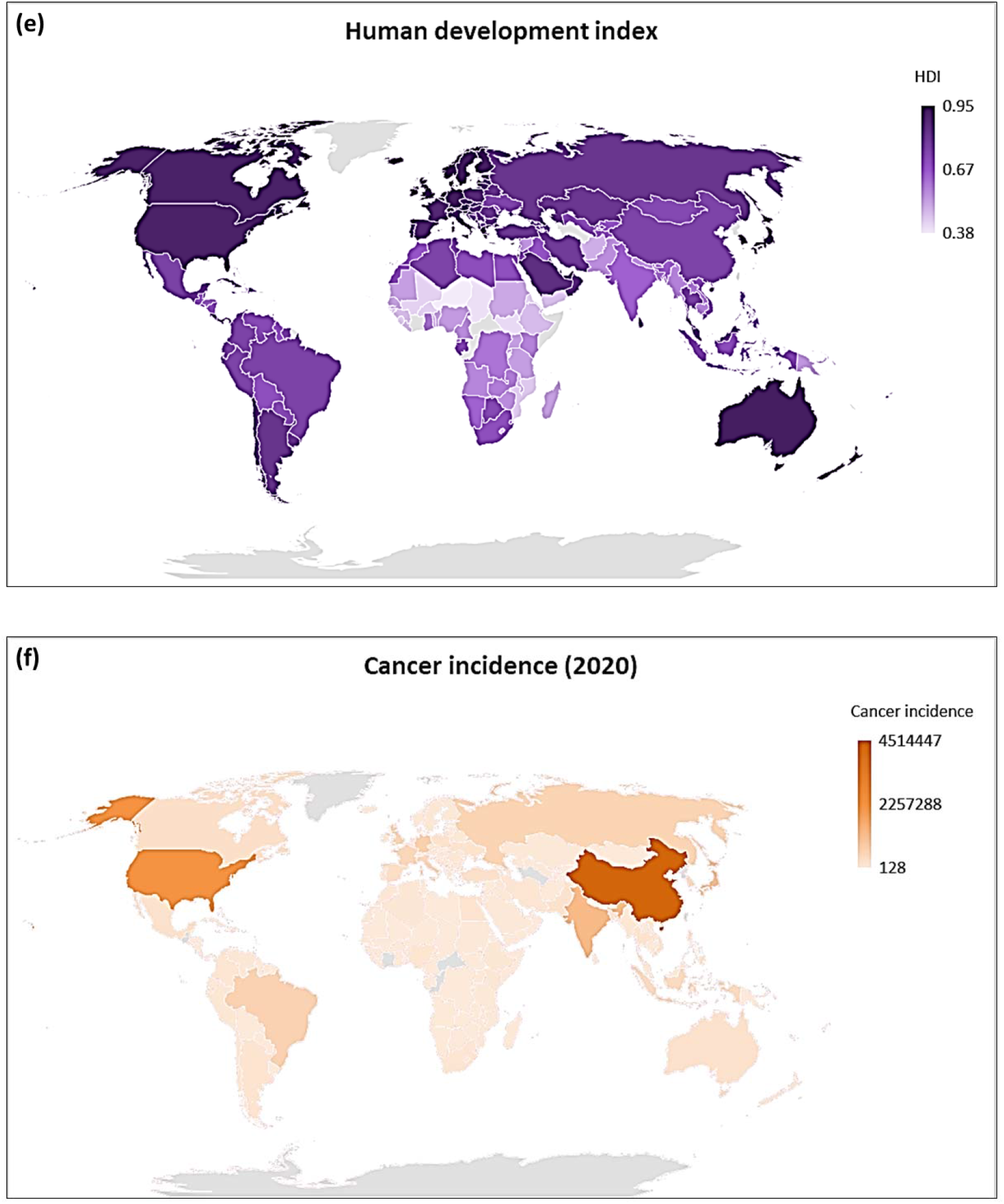
Global distribution map of (a) %case fatality rate due to COVID-19 as reported on July 7, 2020 (b) % BCG coverage (c) % population aged 65+ years (d) Ischemic heart disease/10^5^ population (e) Human development index (f) Cancer incidence (2020).

**Table 1:** Profile of the epidemiological demographic parameters as on July 30, 2020.

| **Epidemiological parameters** | **Number** | **Range** | **Mean ± SD** |
| --- | --- | --- | --- |
| COVID -19 cases / million<sup>a</sup> | 220 | 3.0 – 39340.0 | 2905.1 ± 4849.7 |
| COVID-19 deaths / million<sup>a</sup> | 220 | 0.0 – 1238.0 | 83.51 ± 161.8 |
| COVID -19 case fatality rate<sup>a</sup> (%) | 220 | 0.0 – 28.3 | 3.05 ± 3.5 |
| % BCG vaccination in 1-yr old<sup>b</sup> | 144 | 37.0 – 99.0 | 91.3 ± 10.5 |
| % Population aged 65+ years | 185 | 1.1 – 27.6 | 9.11 ± 6.3 |
| Ischemic heart disease/10<sup>5</sup> population | 182 | 211 – 6769.9 | 1520. 5 ± 1184.2 |
| Hypertensive heart disease/10<sup>5</sup> population | 182 | 11.6 – 252.0 | 70.8 ± 45.3 |
| Cancer incidence | 178 | 128 – 4514447 | 105001.5 ± 399501.9 |
| Diabetes prevalence (20 – 79 years) | 192 | 1.0 – 23.4 | 8.1 ± 4.2 |
| Incidence of malaria/1000 at risk | 103 | 0.0 – 486.0 | 86.1 ± 129.4 |
| Human development index | 180 | 0.377 – 0.954 | 0.715 ± 0.153 |
| Population density/sq.km of land area | 195 | 0.1 – 20777.5 | 373.3 ± 1683.5 |

<sup>a</sup> as on July 30, 2020
<sup>b</sup> in countries with ongoing BCG vaccination programme

### Epidemiological parameters

BCG immunization program for all inhabitants is currently underway in 144 of the 220 countries with %coverage ranging between 37% and 99% (91.3% ± 10.5, median: 95%) (Table 1, Fig. 2b). In 75 countries, either the BCG is advocated in selective high-risk subjects (n=5) or has been suspended (n=21). In the remaining 49 countries, either it was never used or details on the coverage was not available from both WHO and BCG Atlas sites [12, 13]. All these 75 countries were thus categorized in the group of “BCG: not ongoing.” One country not listed in the WHO or BCG World Atlas sites was excluded from computations related to BCG.

The distribution of the remaining epidemiological parameters evaluated were as follows - %population aged ≥65 years (n=185, 1.1% – 27.6%, median: 7.1%), incidences of IHD/10^5^ population (n=182, 211-6769.9, median: 1171), HHD/10^5^ population (n=182, 11.6 – 252, median: 56.3), cancer (n=178, 128 – 4514447, median: 15726), malaria/10^3^ population (n=103, 0.0 – 486, median: 5.1) and %diabetes (n=192, 1.0 – 23.4, median: 7.1%); HDI (n=180, 0.377 – 0.954, median: 0.737) and population density/sq.km per land area (n=195, 0.1 – 20,777.5, median: 90.3) (Table 1).

### Groups based on case-fatality rates

Based on the percentiles of COVID-19 CFR as defined above, 75, 67 and 78 countries were classified into low, moderate and high-risk. Each of the epidemiological variables were estimated for these groups and compared. The variables found to be significant in terms of the COVID-19 CFR were % BCG coverage (p=0.004), %population aged ≥65 years (p=0.001), incidences of IHD/10^5^ population (p=0.016) and cancer (p<0.001) (Table 2, Figs. 2b-e). The incidence of HHD, diabetes, malaria, HDI and population densities were not found to have any significant impact on the countries grouped as per the COVID-19 CFR.

**Table 2:** Impact of demographic variables on the COVID-19 case fatality rates as on July 30, 2020 of 220 countries grouped as low (≤ 1.3%), moderate (1.4% - 3.1%) and high (>3.2%).

| Parameters |  | % COVID-19 case fatality rate |  |  |
| --- | --- | --- | --- | --- |
| | | Low risk<br>($\leq 1.3\%$)<br>(n=75) | Moderate risk<br>($1.4\% - 3.1\%$)<br>(n= 67) | High risk<br>($\geq 3.1\%$)<br>(n=78) |
| % BCG coverage<sup>a</sup> | Number | 74 | 67 | 78 |
| | Mean ± SD | 61.0 ± 45.5 | 73.5 ± 36.5 | 47.8 ± 45.6 |
|  | Median | 99.0 | 92.0 | 66.5 |
|  |  | p = 0.004<sup>j</sup> |  |  |
| Age (65+) years<sup>b</sup> | Number | 58 | 64 | 63 |
| | Mean ± SD | 6.8 ± 4.6 | 8.8 ± 6.1 | 11.6 ± 7.1 |
|  | Median | 4.9 | 6.8 | 10.7 |
|  |  | p = 0.001<sup>j</sup> |  |  |
| IHD<sup>c</sup> | Number | 57 | 62 | 63 |
| | Mean ± SD | 1191.6 ± 832.8 | 1591.1 ± 1413.2 | 1748.7 ± 1159.7 |
|  | Median | 991.3 | 1151.2 | 1497.0 |
|  |  | p = 0.016<sup>j</sup> |  |  |
| Cancer (2020)<sup>d</sup> | Number | 55 | 62 | 61 |
| | Mean ± SD | 21.595.9 ± 41078.3 | 83292.1 ± 203235.7 | 202268.6 ± 640960.9 |
|  | Median | 7707 | 12950 | 34296 |
|  |  | p < 0.001<sup>j</sup> |  |  |
| HDI<sup>e</sup> | Number | 58 | 60 | 62 |
| | Mean ± SD | 0.70 ± 0.1 | 0.69 ± 0.1 | 0.75 ± 0.2 |
|  | Median | 0.72 | 0.72 | 0.76 |
|  |  | p : ns<sup>j</sup> |  |  |
| HHD<sup>f</sup> | Number | 57 | 62 | 63 |
| | Mean ± SD | 66.8 ± 46.7 | 74.6 ± 43.9 | 70.5 ± 45.5 |
|  | Median | 52.4 | 62.3 | 55.1 |
|  |  | p : ns<sup>j</sup> |  |  |
| Diabetes<sup>g</sup> | Number | 63 | 62 | 67 |
| | Mean ± SD | 8.8 ± 4.8 | 7.6 ± 4.1 | 7.7 ± 3.7 |
| | | Low risk<br>($\leq 1.3\%$)<br>(n=75) | Moderate risk<br>(1.4% - 3.1%)<br>(n= 67) | High risk<br>($\geq 3.1\%$)<br>(n=78) |
|  | Median | 7.8 | 7.1 | 6.9 |
|  |  | p : ns<sup>j</sup> |  |  |
| Malaria<sup>h</sup> | Number | 33 | 40 | 30 |
| | Mean ± SD | 94.7 ± 140.2 | 75.5 ± 114.8 | 90.9 ± 138.6 |
|  | Median | 4.2 | 4.4 | 6.1 |
|  |  | p : ns<sup>j</sup> |  |  |
| PD/sq. km<sup>i</sup> | Number | 64 | 64 | 67 |
| | Mean ± SD | 835.9 ± 2887.1 | 152.9 ± 160.9 | 141.8 ± 202.6 |
|  | Median | 97.9 | 84.0 | 83.3 |
|  |  | p : ns<sup>j</sup> |  |  |

<sup>a</sup> BCG vaccine in 1-yr old (2018) (Countries with no ongoing BCG coverage)
<sup>b</sup> %Population aged 65+ years (2018)
<sup>c</sup> Ischemic heart disease (IHD) : Prevalence for both sexes / 10<sup>5</sup> individuals (2015)
<sup>d</sup> Cancer incidence (2020)
<sup>e</sup> HDI: Human development index (2019)
<sup>f</sup> Hypertensive heart disease (HHD) : Prevalence for both sexes / 10<sup>5</sup> individuals (2015)
<sup>g</sup> Prevalence of diabetes in % population in age group 20 – 79 years (2017)
<sup>h</sup> Incidence of malaria (2018)
<sup>i</sup> Population density (PD): People per sq. km of land area (2018)
<sup>j</sup> Kruskal-Wallis test

### Epidemiological parameters vs COVID-19 event rate and case fatality rate

Countries in which the BCG immunization program is ongoing has a significantly lesser event rate compared to those where this not being advocated (0.034 vs 0.020, p<0.001). For the remaining variables, these were divided into two groups based on the corresponding median values. The covariates found to be significantly influencing the COVID-19 event rate were - %population aged ≥65 years, ≤7.1% vs. >7.1% (0.018 vs. 0.028, p<0.001), incidences of IHD/10^5^ population, ≤1171 vs >1171 (0.018 vs. 0.027, p<0.001), cancer incidence, ≤15726 vs. >15726 (0.018 vs. 0.026, p=0.003) and HDI, ≤0.737 vs >0.737, (0.019 vs. 0.026, p=0.003) (Figs. 2b-f). HHD/10^5^ population (≤56.26 vs > 56.26), malaria/10^3^ population (≤5.12 vs >5.12), %diabetes prevalence (≤7.15 vs >7.15) and population density/sq. km (≤90.3 vs >90.3) had no significant impact on the COVID-19 event rate (Fig. 3).

**Fig. 3.**
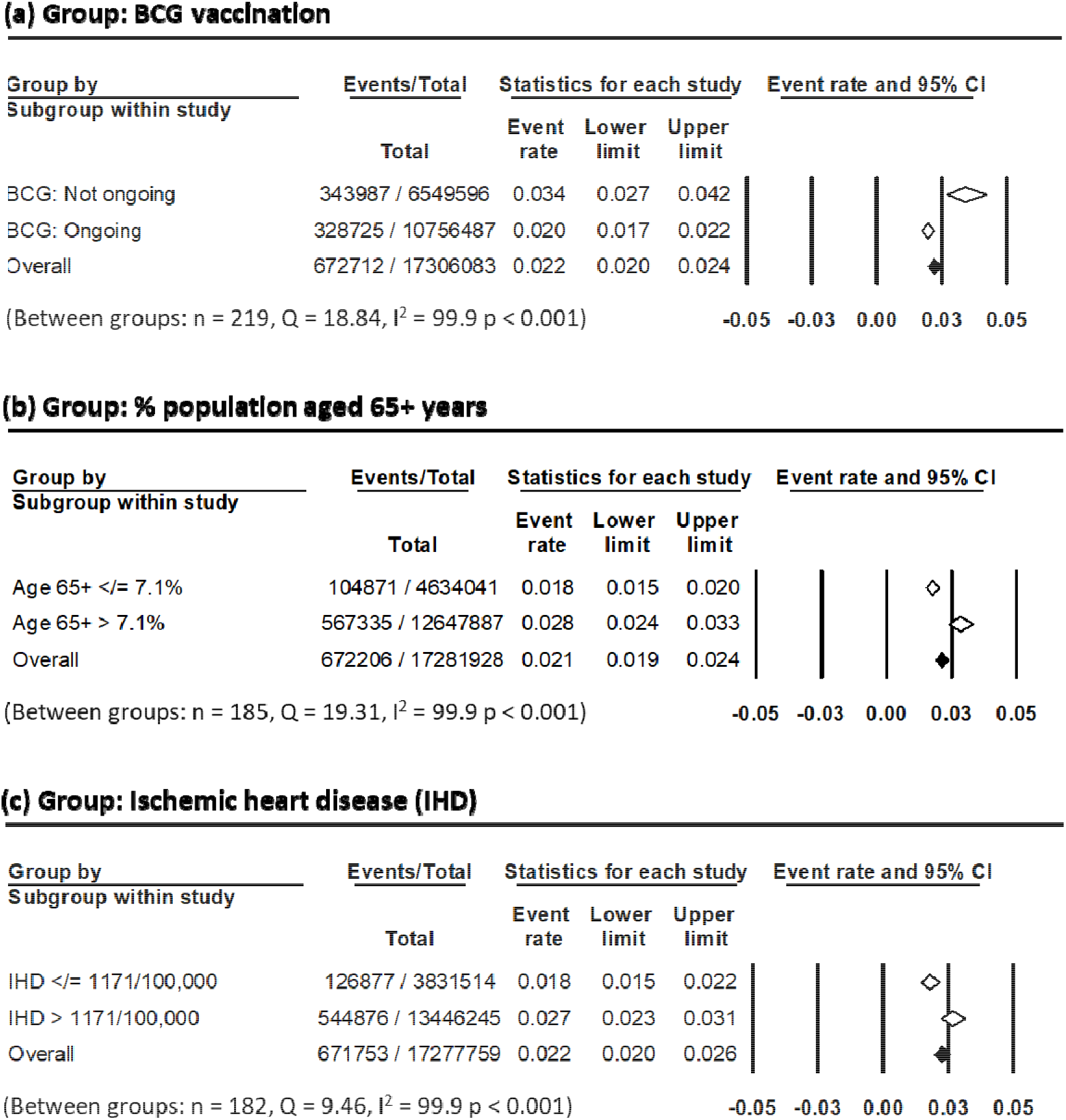
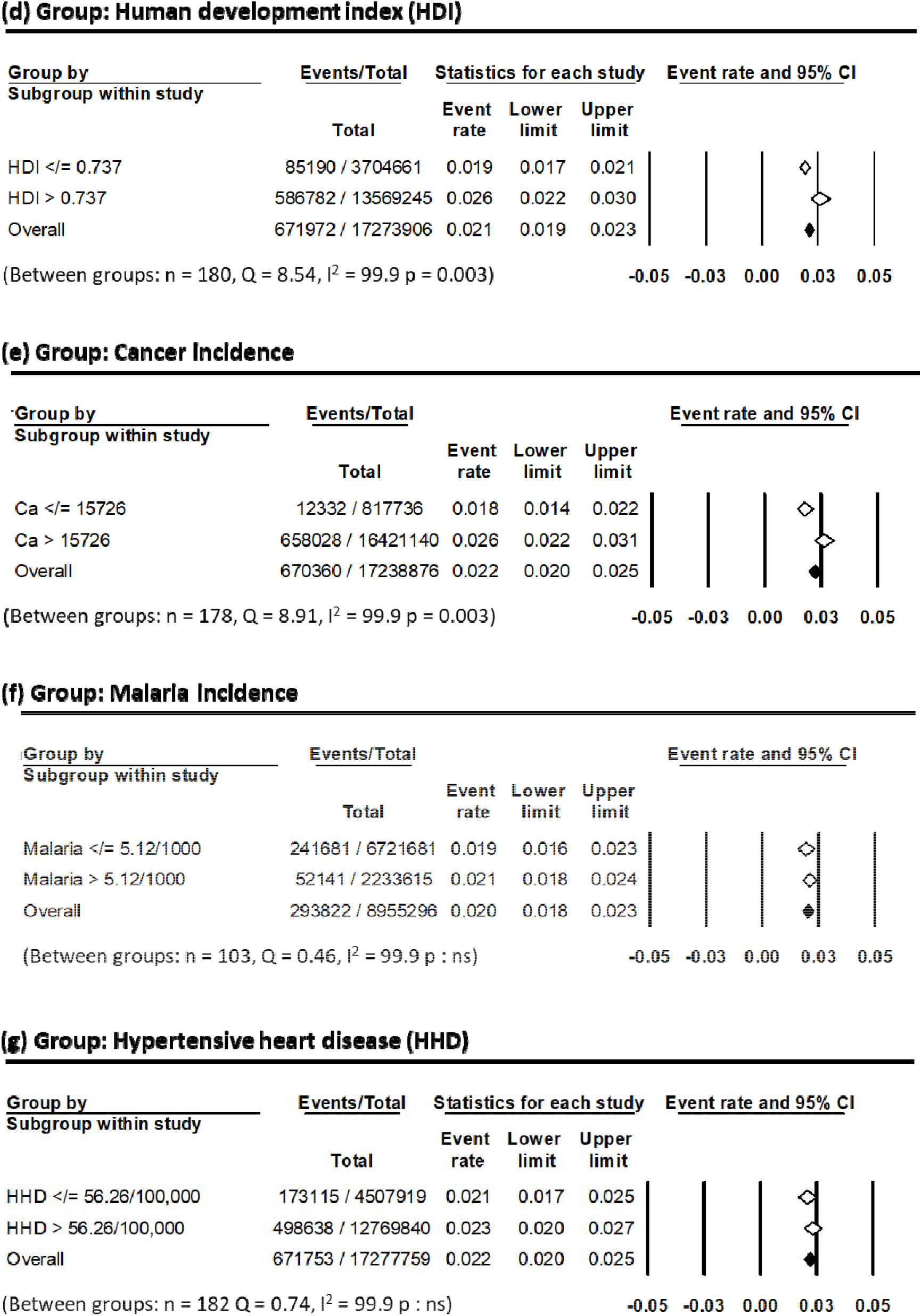
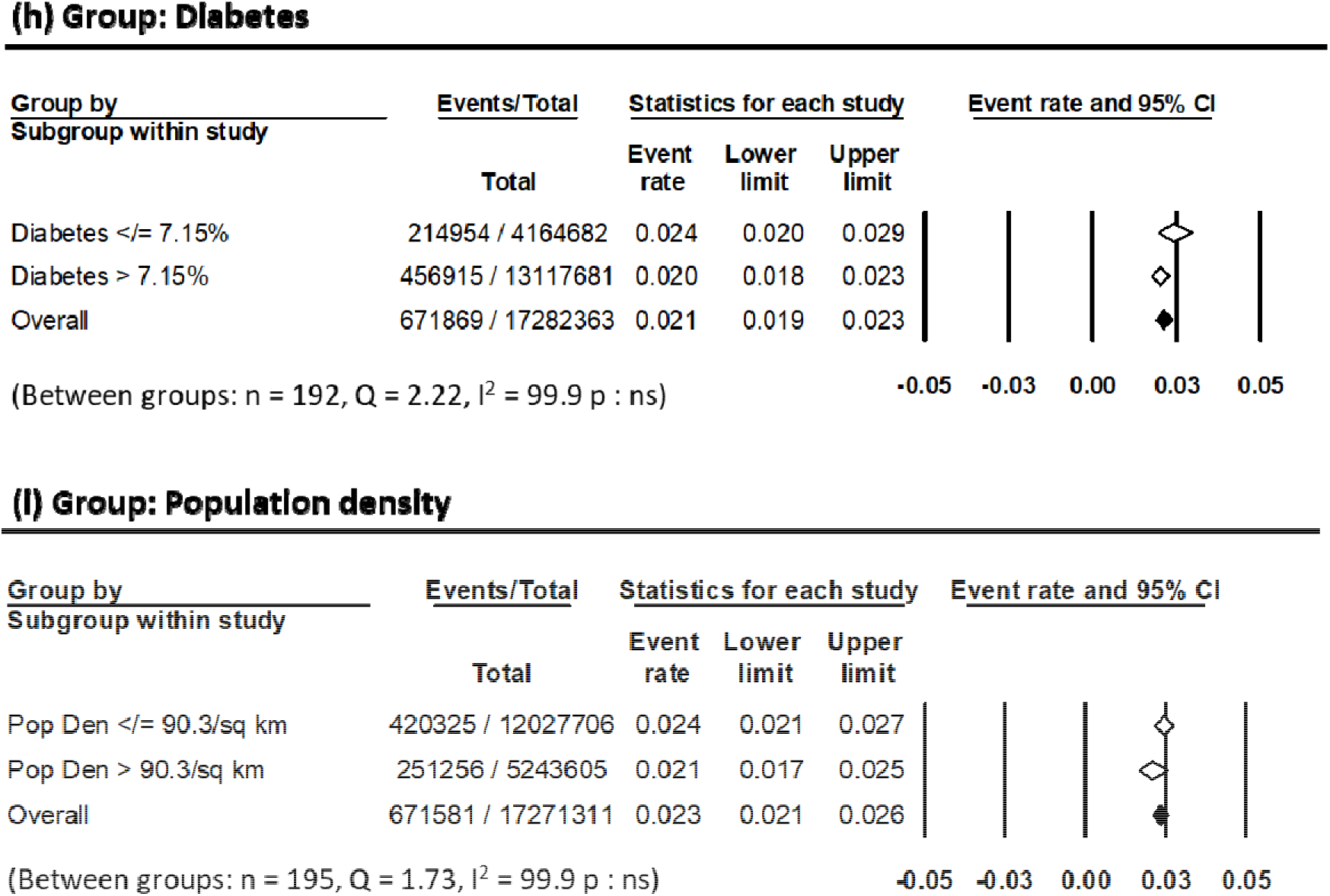
Forest plots for subgroup analysis for covariates for COVID-19 case fatality rate as on July 7, 2020 (a) BCG vaccination in various countries (not ongoing vs ongoing) (b) %Population aged 65+ years (c) Ischemic heart disease/10^5^ population (d) Human development index (e) Cancer incidence (f) Incidence of malaria/1000 at risk (g) Hypertensive heart disease/10^5^ population (h) Diabetes prevalence (20 – 79 years) and (i) Population density/sq.km of land area

The COVID-19 CFR were also compared for the two groups of all the epidemiological parameters. These are summarized as - for BCG, ongoing vs not ongoing (median CFR: 2% vs 2.65%, p=0.04), %population aged ≥65 years, ≤7.1% vs. >7.1% (median CFR: 1.7% vs 2.8%, p=0.004), incidences of IHD/10^5^ population, ≤1171 vs >1171 (median CFR: 1.9% vs 2.5%, p=0.01), cancer incidence, ≤15726 vs. >15726 (median CFR: 1.7% vs 2.8%, p=0.001) and HDI, ≤0.737 vs >0.737, (median CFR: 1.7% vs 2.8%, p=0.002). No significant differences were observed with incidence of HHD, malaria and diabetes and population densities of these countries. Thus the influence of epidemiological covariates on COVID-19 event rates and CFR were in good agreement.

### Impact of BCG on the higher risk subgroups of significant epidemiological parameters

BCG vaccination was found to significantly reduce the COVID-19 event rate for each of the high-risk subgroups of key epidemiological covariates. Thus event rates for BCG vaccination (not ongoing vs. ongoing) for each of the high risk subgroups were - %population of 65+ years >7.1% (0.036 vs 0.023, p =0.008), IHD >1171/10^5^ (0.035 vs 0.022, p=0.004), HDI >0.737 (0.037 vs 0.020, p=0.001), and cancer incidence >15726 (0.043 vs 0.022, p<0.001) (Fig.4).

**Fig. 4.**
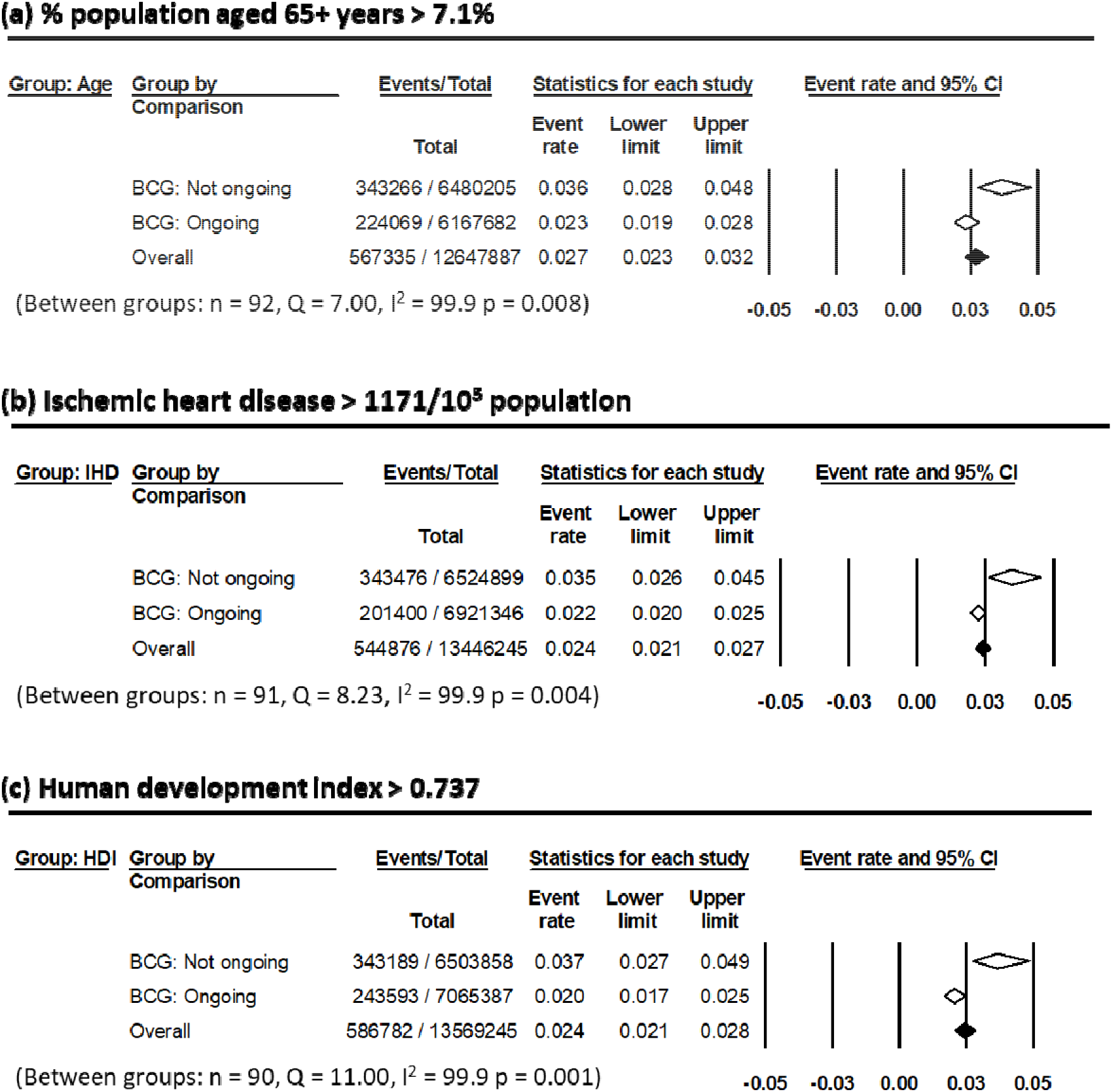
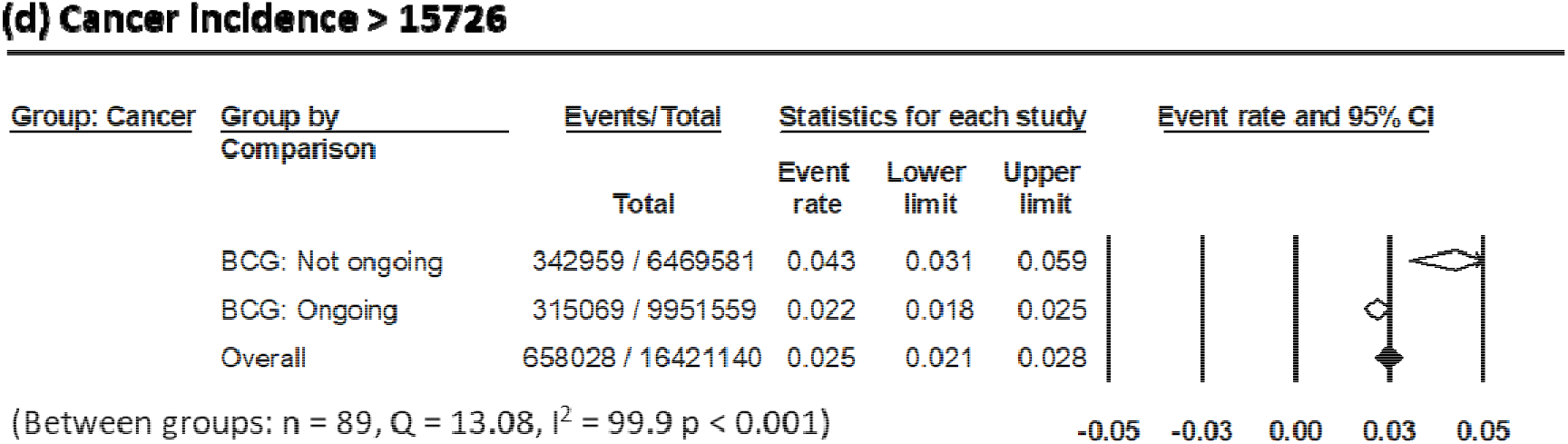
Impact of the none/ suspended vs. ongoing BCG vaccination programme on the COVID-19 case mortality rates for the high risk population (a) % Population aged 65+ years (> 7.1%) (b) Ischemic heart disease/10^5^ population (> 1171/10^5^) (c) Human development index (> 0.737) and (d) Cancer incidence (> 56.25)

### Impact of type of BCG strains on COVID-19 case fatality rate

Of the 144 countries with ongoing BCG vaccination program, the type of strains used was available in 127. Of these, 35 countries advocated early BCG strains while in 27, late BCG strains was used for immunization. Mixed strains was adopted in 65 countries. The COVID-19 event rates computed were 0.018, 0.031 and 0.019 for early, late and mixed strains respectively (p=0.008) (Fig. 5).

**Fig. 5.**
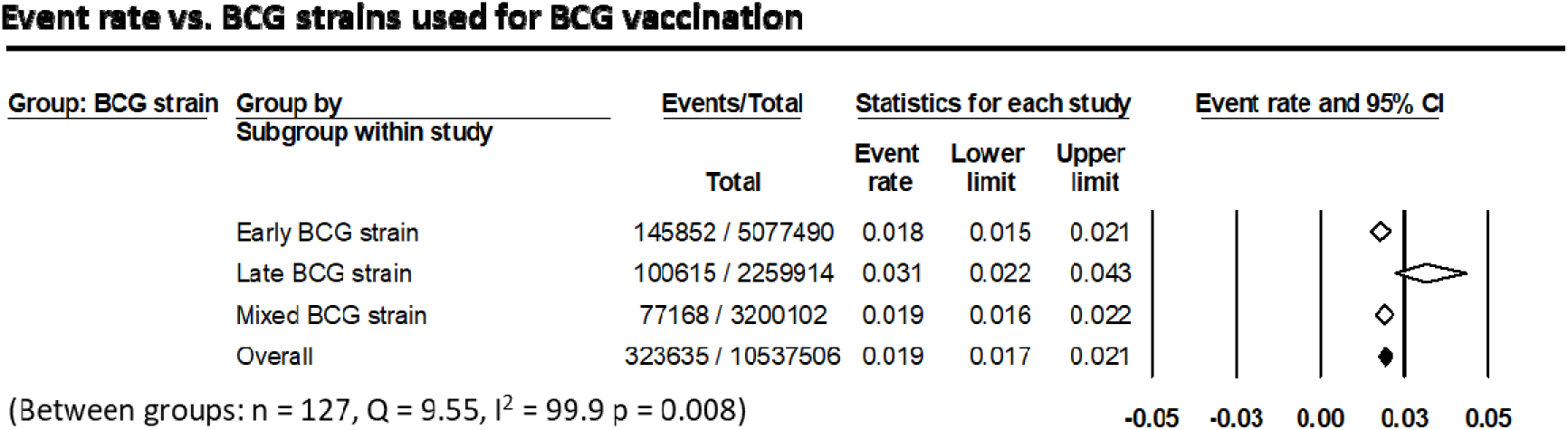
Impact of BCG strains on the COVID-19 case fatality rates in countries with ongoing BCG vaccination. This represent 127 of the 144 countries for which the BCG vaccination strains were available.

## Discussion

The ferocity of development of COVID-19 pandemic has led to an unprecedented thrust to accept the challenge and explore various options to help lessen its global impact on health, economy and society. While the search continues to find the most optimum therapeutic combination, the quest for a specific vaccine directed towards SARS-CoV-2 remains the ultimate focus of the current global research. Concurrently, a number of reports have looked into the various demographical factors that appear to influence the prevalence and mortality of COVID-19 [1-9]. However, even if these could attribute towards the prevalence and mortality of COVID-19, the ability to effectively modify these variables through prospective interventions are limited.

One of the variables, %BCG coverage has widely varied across the 220 countries facing the pandemic [12, 13]. BCG provides heterologous immunoprotection against a wide range of microbes including respiratory and yellow fever viruses [25-28]. The nonspecific protection offered by BCG vaccine could be due to similarities between the viral and the BCG antigens, antigen independent activation of the bystander B-and T-cells or long-term activation and reprogramming of the innate immune cells [25]. This is proposed as development of “trained immunity”[29]. This immunity is independent of T-and B-cell memory and provides a nonspecific cross-protection contributed by the innate immune cells - macrophages and natural killer cells. An excellent summary of the possible mechanisms of BCG induced trained and its likely impact on COVID-19 has been recently outlined by O’Neill and Netea [7].

In view of the gross heterogeneity in the reported COVID-19 case fatality, the present study had evaluated the impact of various epidemiological parameters and their interactions on the outcomes covering a population of over 7.76 billion in 220 countries. Population ≥65 years and associated comorbidities like IHD and cancer have shown to negatively influence the outcomes (Table 2 and Fig.3). The impact of HDI was not evident on the three group classifications based on COVID-19 CFR (Table 2). However, countries with higher HDI showed a significantly higher event rate (HDI: ≤0.737 vs >0.737, event rate 0.019 vs. 0.026, p=0.003) (Fig.3). This could be perhaps attributed to the fact that in most HDI countries had no ongoing BCG immunization for its population (Spearman correlation: −0.162, p=0.03). Higher prevalence of diabetes, malaria and HHD along with a higher population density were not significantly associated with higher COVID-19 CFR in their group classification (Table 2) nor on the event rate (Fig.3).

BCG vaccination has shown to mitigate the influence of BCG on COVID-19 [8, 21, 30, 31]. However, these studies have usually been reported looking at BCG only. Since we expect that other epidemiological parameters could also have an influence on the COVID-CFR, we examined the most relevant parameters that could be accessed in the public domains and thereby identify the high-risk population who could be most vulnerable and need protection. A reduction in the event rate from 0.034 with no active BCG vaccination to 0.020 with ongoing BCG immunization program represents a 41.1% reduction in the event rate (Fig. 3a). Further BCG vaccination appears to significantly reduce the COVID-19 event rate in all high-risk subgroups of population ≥65 years (↓36.1%), higher incidences of IHD (↓37.1%), cancer (↓48.8%) and countries with higher HDI (↓45.9%) (Fig. 4).

The impact of immunogenicity induced by BCG could be dependent on the type of strains. The late strains are deficient in their ability to produce cell wall methoxymycolic acid which constitute an key group of ligands proficient in inducing trained immunity [21], Our results lend support to this hypothesis as the event rate was significantly lower in countries using early strain BCG (0.18) (Fig.5). The event rate with late BCG strains of 0.031 was nearly the same as those countries without BCG vaccination at 0.034 (Figs. 3a and 5). In contrast, those with early and mixed strains, the event rates were nearly similar at 0.018 and 0.019 respectively.

As on July 30, 2020, of the 10 countries with the worst COVID-19 CFR ranging from 11.1% to 28.3%, seven had no ongoing BCG vaccination program. Of the remaining three countries, two used late strains of BCG while one had mixed BCG strain. Even though the event rate for mixed BCG strain was nearly similar to those with early strain (0.019 vs 0.018, Fig.5), the %BCG coverage in this country (Yemen) was grossly inadequate at just 64%. Thus, even though other key prognostic epidemiological covariates like population aged 65 years and above, incidence of IHD and cancer were favorable, but inadequate BCG vaccination coverage could have been one of the key contributing factors towards the highest recorded COVID CFR at 28.3% in Yemen.

The duration of lasting of the BCG induced trained immunity is of significance as BCG vaccination are usually given within 1-year of birth. Netea et al [28, 29] had demonstrated that the trained immunity status was maintained at least for one year which was the maximum observed time point in their report. A recent report [21] from 13 European countries who had suspended their BCG vaccination program more than two decades ago, suggests that the BCG induced heterologous non-specific protective effect could even last for 20 years or more.

BCG vaccination may not be expected to reduce the number of positive cases, most of whom could recover through the “trained immunity” imparted by BCG. However as evident from this analysis and supported by other studies, BCG could reduce the fatal outcomes [8, 21, 30, 31]. One could well argue that the results of this analysis would also depend on the number of tests performed. However, the testing would certainly influence the cases/million and deaths/million population estimates. Thus, to minimize the impact of testing, the impact of BCG at the fatality in those tested positive for COVID-19 was evaluated in this study. Even though a widespread testing would be desirable, one has to accept that the testing protocols adopted by countries are varied and depends on their individual policies, availability and logistics of carrying out these tests. These tests have not been used in most countries as a screening procedure for all subjects. However, tests would have been conducted in most of the suspected, symptomatic, contacts of positive patients, high-risk individuals and those subjected to compulsory screening (travelers etc.). Even with all the inherent limitations as discussed above, the present data should be construed as a reasonably adequate sample of more than 332.84 million test reports obtained globally reported as on July 30, 2020.

Despite these constraints, the evidence of a likely protective effect of BCG vaccination on COVID-19 is apparent. It would be imperative to examine the effects of BCG vaccinations through well designed randomized clinical trials. Presently, four phase III randomized clinical trials have been initiated, one each in Netherlands (NCT04328441), Australia (NCT04327206), USA (NCT04348370) and South Africa (NCT04379336) [32], These are being conducted in healthcare workers randomized between BCG and placebo injection of normal saline. The outcomes from these studies would confirm the observational findings of a potential benefit of BCG vaccination against COVID-19.

The data from 220 countries and dependent territories represents 7.76 billion inhabitants globally [33]. Even though this is an observational study, the alleviating impact of BCG vaccination on COVID-19 cannot just be overlooked while we are still investigating to arrive at the optimum pharmacotherapy and racing against time towards developing a specific vaccine. However, the results of this analysis should not be construed as a gateway for advocating BCG vaccination for all. Nor should it deliver a false sense of security and complacency to those who have been previously vaccinated. It does not in any way dilute the usual precautions to be adopted for preventing COVID-19. Nonetheless, it does support the hypothesis of “trained immunity” with BCG vaccination. Pending the results of the ongoing phase III clinical trials, the study could be considered as suggestive of a cost-effective prophylaxis by BCG vaccination especially for high-risk population. In countries which already have an ongoing BCG vaccination during neonatal period, the effectivity of a booster dose of BCG selective for high-risk population needs careful evaluation and is beyond the scope of this manuscript.

The conclusions derived from this study could be further examined through study of individual cases and needs joint efforts from various national and international agencies. This study, however provides a pragmatic rationale for BCG vaccination with early strains in high-risk individuals to reduce the COVID-19 case fatality to bridge the gap till an effective vaccine specifically against SARS-CoV-2 is freely available globally.

## Data Availability

Data used is available in the Supplementary Table 1. The data have been retrieved from the public domains of the websites as mentioned in the text. All these data are freely available online.

## Acknowledgement

We acknowledge the inputs from Prof. M. Borenstein on the analysis and Dr. Indranil Pan for his support and guidance regarding programming in Python for data extraction and analysis. Discussions with Prof. C.M. Pandey is also gratefully acknowledged.

## Supporting information

**S1 Table:** COVID-19 status shown as cases/million, deaths/million, case fatality rate as on July 30, 2020 for % BCG vaccination in 1-yr old, % population aged 65+ years, human development index (HDI), ischemic heart disease (IHD)/10^5^ population, hypertensive heart disease (HHD)/10^5^ population, cancer incidence, incidence of malaria/1000 at risk, diabetes prevalence (20 - 79 years), population density (PD)/sq.km of land area and BCG strain

## Authors contributions

**Conceptualization:** Niloy R. Datta

**Data curation:** Sneha Datta and Niloy R. Datta

**Formal analysis:** Niloy R. Datta

**Supervision:** Niloy R. Datta

**Writing original draft:** Niloy R. Datta and Sneha Datta

**Writing-reviewing and editing:** Niloy R. Datta and Sneha Datta

